# Effect of antibacterial mouthwash on NO bioavailability and muscle contractile function in young healthy men and women

**DOI:** 10.1101/2024.05.21.24307698

**Authors:** Edgar J. Gallardo, William S. Zoughaib, Ahaan Singhal, Richard L. Hoffman, Andrew R. Coggan

## Abstract

The use of antiseptic mouthwash is widespread due to its oral health benefits. However, its impact on systemic physiological processes, particularly nitric oxide (NO) bioavailability and muscle contractility, is not fully understood.

**PURPOSE:** To determine the effects of cetylpyridinium-based (antibacterial) versus sodium chloride (NaCl) -based (control) mouthwashes on salivary and breath NO markers and muscle contractile function in healthy young adults.

**METHODS:** Twenty-six participants (n=13/group) completed a randomized, parallel-arm, blinded trial, comparing the effects of the two mouthwashes before and after 7 d of treatment. NO bioavailability was assessed via measurement of nitrate (NO_3_^-^), nitrite (NO_2_^-^), and cyclic guanyl monophosphate (cGMP) concentrations in saliva and the level of NO in breath. The contractile function of the knee extensor muscles was determined via isokinetic dynamometry.

**RESULTS:** No significant changes in salivary NO_3_^-^, NO_2_^-^, or cGMP or in breath NO were observed in response to either treatment. However, cetylpyridinium-based mouthwash reduced the percentage of NO_3_^-^ in saliva (16.9±10.5% vs. 24.9±13.4%; p=0.0036), supporting compliance with the intervention. Peak torque at velocities of 0-6.28 rad/s was unaffected by mouthwash use. Calculated maximal knee extensor velocity (Vmax) and power (Pmax) were therefore also unchanged.

**CONCLUSION:** Cetylpyridinium-containing mouthwash inhibits reduction of NO_3_^-^ to NO_2_^-^ in the oral cavity but does not significantly diminish overall NO bioavailability or impair muscle contractile function in healthy young men and women.

**ClinicalTrials.gov Registration Number:** NCT04095442 (registered 09/19/2019)

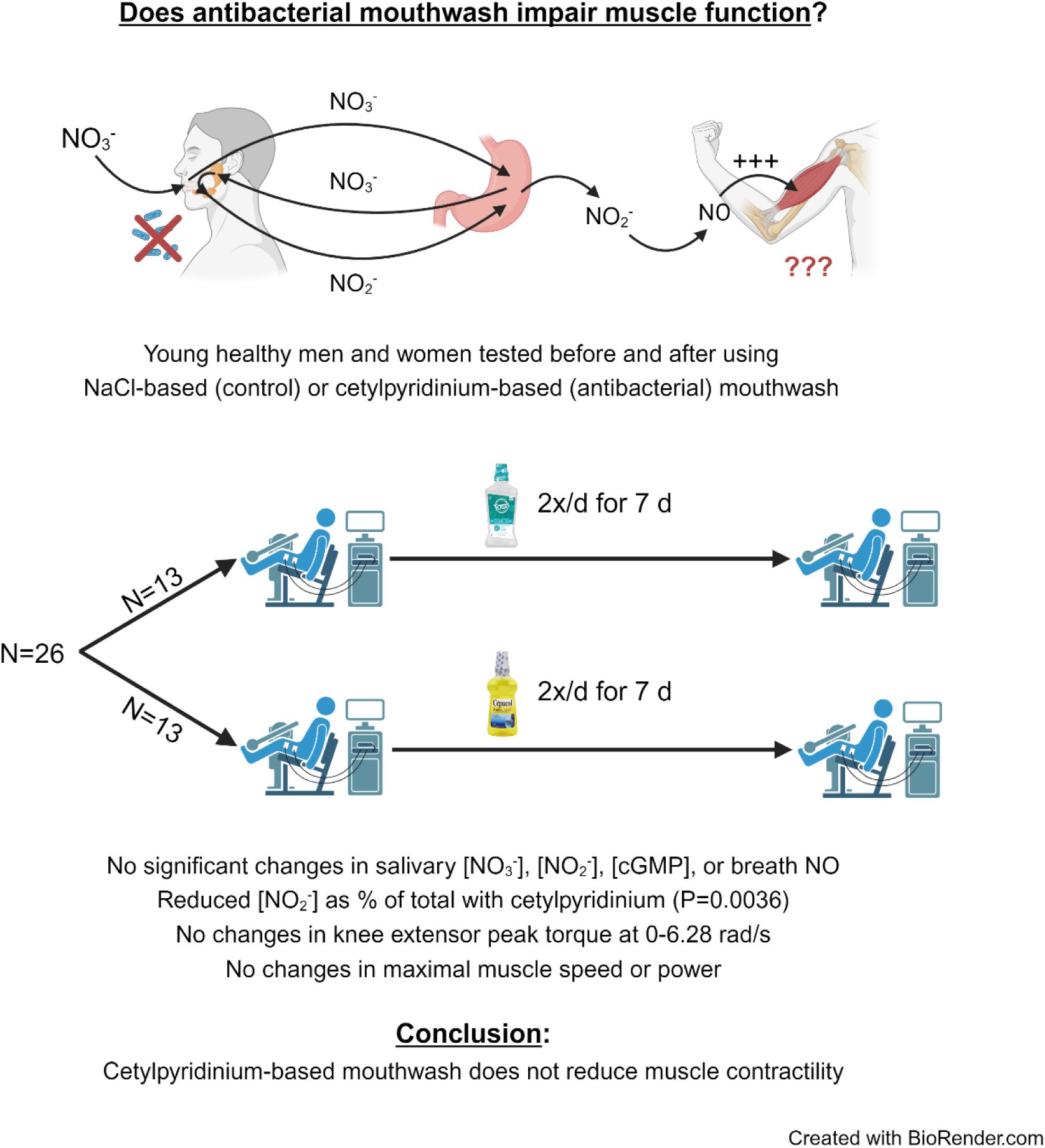

## Introduction

Approximately $7B worth of antiseptic/antibacterial mouthwash is sold globally each year (Grand View Research, 2024), with roughly one-third to one-half of adults in many countries using such products on a regular basis (Górska and Górski 2018; McFarlane et al. 2018; Luzzi et al. 2020). Antiseptic/antibacterial mouthwash use can improve oral health, by diminishing plaque formation and reducing gingivitis (Rajendiran et al. 2021). However, such products may also have detrimental effects. In particular, prior research has demonstrated that strong antibacterial mouthwash can disrupt the nitrate (NO_3_^-^) -reducing bacteria normally found in the mouth (Mitsui and Harasawa 2017; Tribble et al. 2019; Ashworth et al. 2019; Bescos et al. 2020). These facultative anaerobes reduce salivary NO_3_^-^ to nitrite (NO_2_^-^), which after being swallowed and absorbed into the circulation can be further reduced by, e.g., deoxyhemoglobin, to form nitric oxide (NO) (Lundberg and Weitzberg 2010; Amdahl, DeMartino, and Gladwin 2020). NO is a key signaling molecule involved in a variety of physiological responses, including, but not limited to, the regulation of blood flow/pressure. As a result, interruption of the bacteria-dependent enterosalivary pathway of NO production via twice-daily use of chlorhexidine-containing mouthwash for 3-7 d has been reported to decrease plasma NO_2_^-^ levels by 15-25% and increase systolic blood pressure by 2-3 mmHg (Kapil et al. 2012; Bondonno et al. 2015). The latter change is potentially clinically significant, as it would raise the chances of dying of ischemic heart disease and stroke by 7 and 10%, respectively (Lewington et al. 2002). In fact, frequent use of over-the-counter mouthwash has been associated with the development of hypertension (Joshipura et al. 2020), as well as pre-diabetes/diabetes (Joshipura et al. 2017). Chronic mouthwash use therefore may have unintended negative consequences.

In addition to influencing blood flow/pressure, NO is involved in the regulation of numerous other physiological functions, e.g., neural transmission, immune function, mitochondrial respiration, etc. This includes regulation of the contractility of skeletal muscle (Maréchal and Gaily 1999). In this regard, numerous studies in recent years have demonstrated that acute ingestion of NO_3_^-^-rich beetroot juice can enhance muscle contractile function in a wide variety of individuals (cf. Coggan et al. 2021 for review). The exact mechanism responsible for this beneficial effect is still unclear, but it has been hypothesized to be the result of changes in Ca^2+^ release and/or sensitivity due to NO-stimulated nitros(yl)ation of the ryanodine receptor or phosphorylation of the regulatory light chain of myosin (Coggan and Peterson 2018). Indeed, muscle function seem to be more sensitive to alterations in NO availability than blood pressure, as we have routinely observed improvements in the former in response to dietary NO_3_^-^ supplementation even in the absence of changes in the latter (Coggan et al. 2015, 2020; Gallardo et al. 2021; Zoughaib et al. 2023). It is not known, however, whether a *reduction* in NO bioavailability resulting from use of antibacterial mouthwash can *inhibit* muscle function. Such knowledge is obviously relevant not only in the context of athletic performance but also to clinical populations (e.g., the elderly) in whom baseline muscle strength, speed, and power are impaired, thus limiting activities of daily living and increasing the risk of falls.

The purpose of the present study was therefore to determine the effects of antibacterial mouthwash on muscle contractility in healthy young men and women. We hypothesized that this would result in a significant decrease in NO bioavailability, as reflected by salivary NO_3_^-^, NO_2_^-^, and cGMP concentrations and/or breath NO levels, which in turn would result in a decline in muscle contractile function.

## Methods

### Participants

Potential participants were recruited via word-of-mouth and flyers placed around the university campus. Exclusion criteria were age <18 or >30 y; current use of mouthwash, antibiotics, or tobacco products; resting blood pressure > 140/90; an answer of yes to any of the seven general health questions of the Physical Activity Readiness Questionnaire (PAR-Q); or inability to provide informed consent. All other persons were included. After screening of 32 individuals, 16 men and 10 women ultimately completed the study. Each provided written, informed consent, and the study protocol was approved by the Human Subjects Office at Indiana University.

### Experimental design and protocol

After an initial screening visit, which included practicing the isokinetic dynamometer testing protocol described below, participants were randomized (via randomization.com) in blocks of four to one of two treatments. One group of participants was studied before and after using Cepacol® mouthwash (Reckitt Benckiser, Parsippany, NJ) for 7 d. This product was chosen because it is the strongest non-prescription antibacterial mouthwash available in the United States (Jenkins et al. 1994). As a comparison/control, a second group was studied before and after using Tom’s of Maine® Sea Salt Natural Mouthwash (Kennebunk, ME) for 7 d. Based on previous research (Hoover et al. 2017), this product was not expected to have any significant effects on the oral microbiota and hence NO production. These two treatment groups will henceforth be referred to by the active ingredients of the two products, i.e., cetylpyridinium and sodium chloride (NaCl), respectively. Characteristics of the participants in each group are shown in Table 1.

**Table 1.**
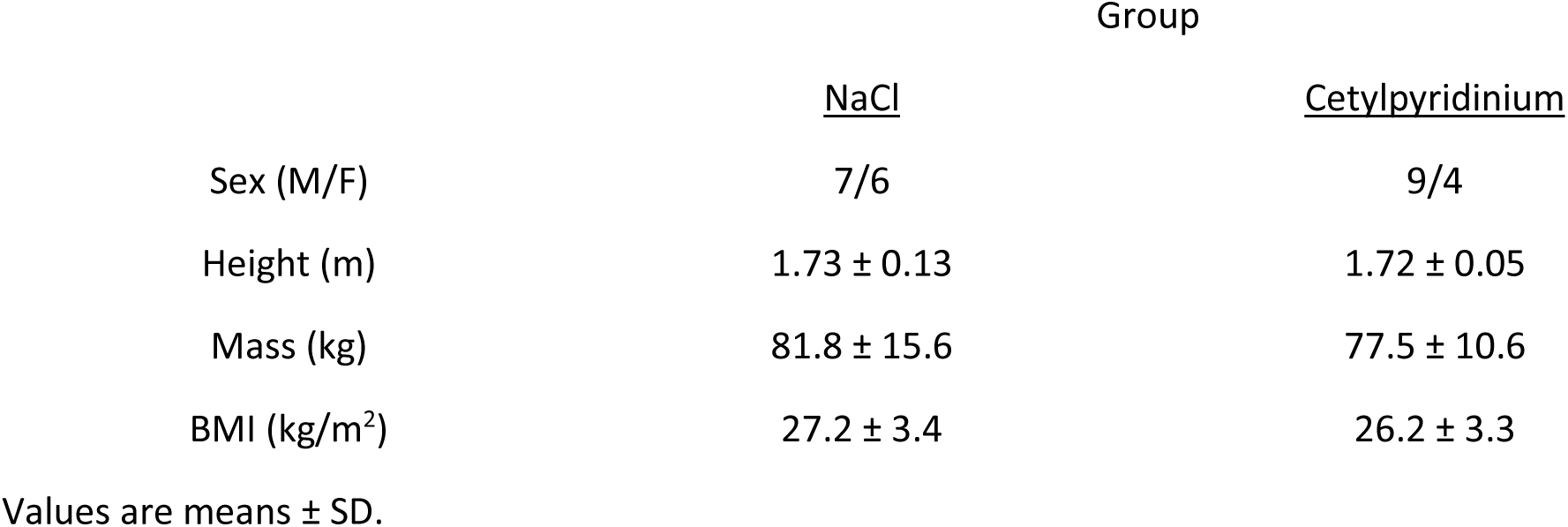
Participant characteristics.

|  | Group |  |
| --- | --- | --- |
|  | <u>NaCl</u> | <u>Cetylpyridinium</u> |
| Sex (M/F) | 7/6 | 9/4 |
| Height (m) | 1.73 ± 0.13 | 1.72 ± 0.05 |
| Mass (kg) | 81.8 ± 15.6 | 77.5 ± 10.6 |
| BMI (kg/m<sup>2</sup>) | 27.2 ± 3.4 | 26.2 ± 3.3 |

Values are means ± SD.

Participants in both groups were instructed to refrain from exercising and to avoid alcohol, caffeine, chewing gum, and foods high in NO_3_^-^ (e.g., spinach, beets, collard greens) for 24 h prior to study visits. Upon reporting to the Exercise Physiology Laboratory, the level of NO in the participant’s breath was first measured using a portable electrochemical analyzer (NIOX VERO®, Circassia Pharmaceuticals, Mooresville, NC). (Note that due to equipment obsolescence, these measures were only obtained in the first eight participants in each group.) A saliva sample was then collected using a commercial collection kit (Salivette, Sarstedt, Newton, NC). This sample was immediately centrifuged for 2 min at 1000 g and 4° C and then frozen at -20° C until subsequently analyzed for NO_3_^-^ and NO_2_^-^ concentrations using a dedicated high performance liquid chromatography system (ENO-30, Amuza, San Diego, CA). Salivary cCGMP levels (which are highly correlated with plasma concentrations (Gauquelin et al. 1992)) were also determined in these samples using a commercial ELISA kit (Item number 581021, Cayman Chemical, Ann Arbor, MI).

The contractile properties of the quadriceps muscle group were next determined using an isokinetic dynamometer (Biodex System 4 Pro, Biodex Medical Systems, Shirley, NY) as previously described (Coggan et al. 2015, 2020; Zoughaib et al. 2023). Briefly, participants performed three maximal knee extensions at angular velocities of 0, 1.57, 3.14, 4.71, and 6.28 rad/s (0, 90, 180, 270, and 360 °/s), with 2 min of rest allowed between each set of contractions. The torque data were windowed and smoothed using the manufacturer’s software, after which the highest torque generated at each velocity was used to calculate peak power at that velocity. The peak power–velocity data were then fit with a parabolic function to determine the maximal velocity (Vmax) and power (Pmax) of knee extension.

Upon completion of the baseline visit, participants were provided with a 7 d supply (i.e., one bottle) of their assigned mouthwash in a brown paper bag (to conceal the assignment from the investigators). They were instructed to rinse their mouth as directed on the product’s packaging for 30 s twice per day for 7 d and to record the usage of the mouthwash on a provided form. They were also instructed to not use any other mouthwash products during this period. Note that although the two products differed significantly in terms of packaging, color, taste, etc., the participants were simply told that the purpose of the study was to compare two different mouthwashes, with no expectation as to the outcome, and were kept unaware of the other (i.e., non-assigned) product being used in the study. After 7 d, they returned to the Exercise Physiology Laboratory where the above measurements were repeated. *Statistical analyses:* Statistical analyses were performed using GraphPad Prism version 10.2.2 (GraphPad Software, La Jolla, CA). Normality of data distribution was tested using the D’Agostino-Pearson omnibus test. Salivary NO_3_^-^, NO_2_^-^, and cGMP concentrations (the latter after log normalization), breath NO levels, and Vmax and Pmax were analyzed using two-way ANOVAs, with treatment as a between-subject factor and time as a within-subject factor. Isometric and isokinetic knee extensor peak torque data were analyzed using a three-way ANOVA, with treatment as a between-subject factor and velocity and time as within-subject factors. Post-hoc testing was performed using the Holm-Šidák multiple comparison procedure. P<0.05 was considered significant.

## Results

### Markers of NO bioavailability

No significant changes were observed in salivary NO_3_^-^ or NO_2_^-^ concentrations or in their sum in either group (Figs. 1A-C). There were also no significant changes in salivary cGMP concentrations or in breath NO levels (the latter measured in only n=8/group) (Figs 1E, 1F). There was, however, a significant reduction in the relative abundance of NO_2_^-^ in the cetylpyridinium group, which decreased 24.9±13.4% to 16.9±10.5% of the total salivary NO_3_^-^ plus NO_2_^-^ concentration (P=0.0036) (Fig. 1D).

**Fig. 1.**
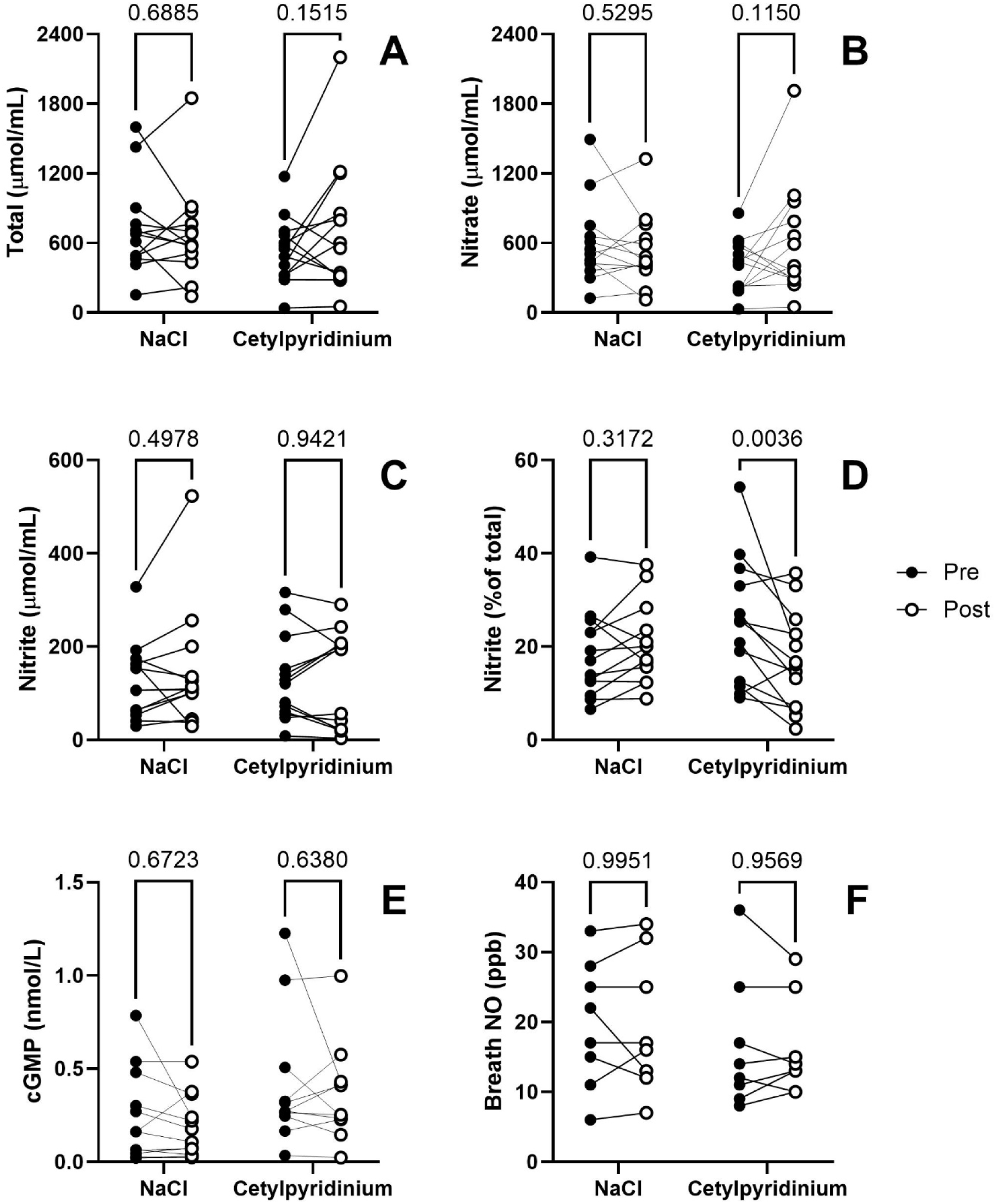
Effect of NaCl- or cetylpyridinium-containing mouthwash on markers of NO bioavailability

### Muscle contractile function

Although there was a highly significant (i.e., P<0.001) effect of velocity on knee extensor peak torque, the effects of time (P=0.4725) and treatment (P=0.5607) were not significant, nor were there any significant interaction effects (Table 2). Vmax (Fig. 2A) and Pmax (Fig. 2B) therefore also did not differ by time or treatment.

**Fig. 2.**
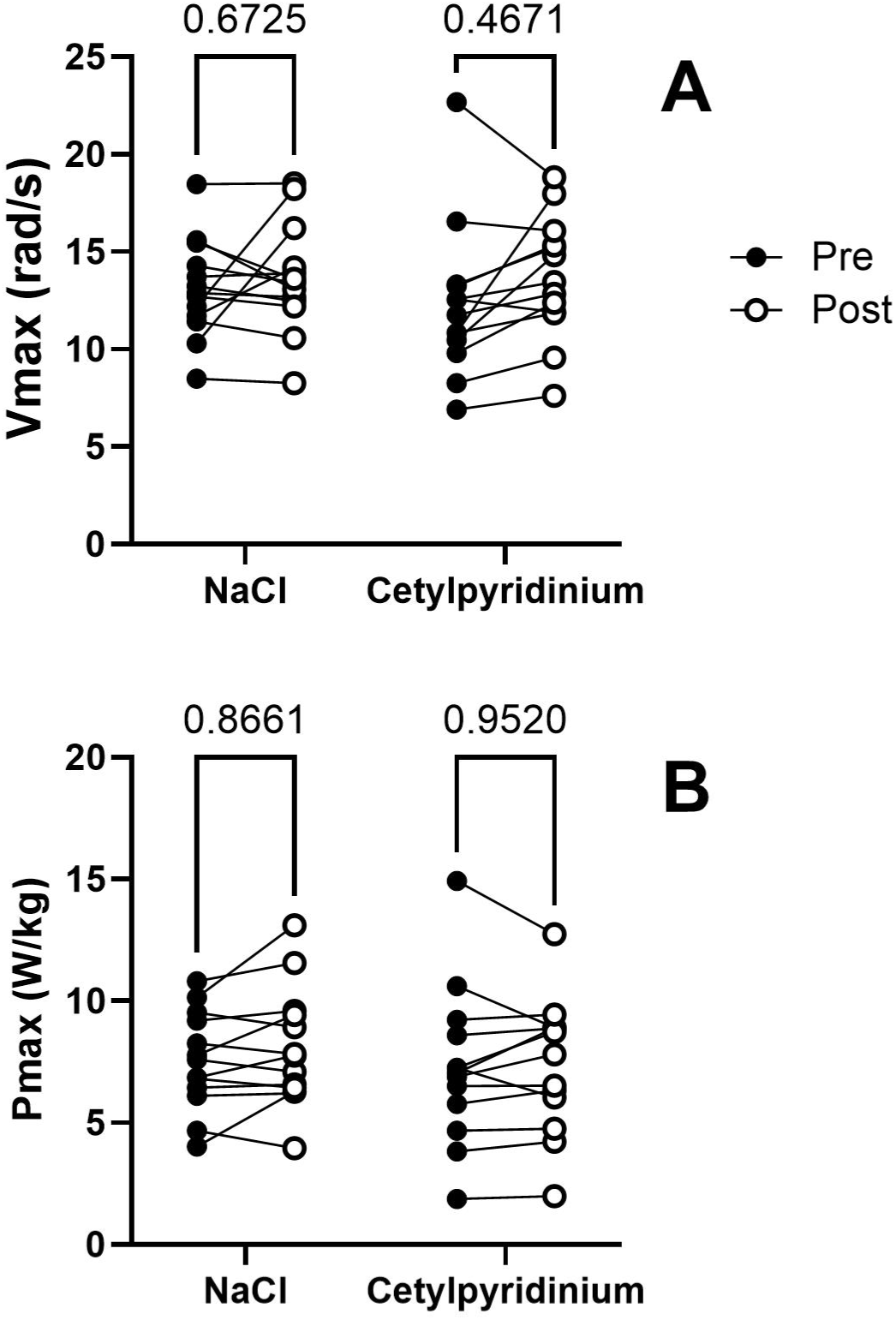
Effect of NaCl- or cetylpyridinium-containing mouthwash on maximal knee extensor velocity (Vmax) and power (Pmax)

**Table 2.** Effect of NaCl- or cetylpyridinium-containing mouthwash on the maximal isometric/isokinetic torque of the knee extensors.

| Velocity<br>(rad/s) | Group |  |  |  |
| --- | --- | --- | --- | --- |
|  | NaCl<br><u>pre</u> | NaCl<br><u>post</u> | Cetylpyridinium<br><u>pre</u> | Cetylpyridinium<br><u>post</u> |
| 0 | 2.63 ± 0.53 | 2.69 ± 0.53 | 2.48 ± 0.86 | 2.49 ± 0.87 |
| 1.57 | 2.14 ± 0.48 | 2.22 ± 0.48 | 2.14 ± 0.66 | 2.09 ± 0.66 |
| 3.14 | 1.75 ± 0.44 | 1.78 ± 0.37 | 1.73 ± 0.62 | 1.63 ± 0.61 |
| 4.71 | 1.43 ± 0.37 | 1.48 ± 0.38 | 1.35 ± 0.50 | 1.34 ± 0.49 |
| 6.28 | 1.17 ± 0.32 | 1.24 ± 0.38 | 1.07 ± 0.47 | 1.13 ± 0.42 |

Values are means ± SD in Nm/kg. No significant treatment effects were observed.

## Discussion

The purpose of this investigation was to determine whether use of antibacterial mouthwash would reduce NO production via the enterosalivary pathway and thus impair muscle contractility in young healthy men and women. Contrary to this hypothesis, we found no significant changes in salivary NO_3_^-^, NO_2_^-^, or cGMP concentrations, in breath NO levels, or in the maximal voluntary isometric/isokinetic torque or speed or power of knee extension. This is the first study to specifically examine the effects of mouthwash use on muscle contractile function or, to our knowledge, any type of human exercise performance.

There are several possible explanations for these negative findings. The first is simply that the participants failed to utilize the provided mouthwash as instructed. However, review of the logs maintained by the participants indicated excellent compliance, averaging 96±7% and 97±4% in the cetylpyridinium and NaCl groups, respectively. Furthermore, as expected/intended the relative abundance of NO_2_^-^ in saliva was significantly reduced in the cetylpyridinium group, as discussed in greater detail below. Thus, while participant non-compliance cannot be completely excluded as a factor, this explanation seems unlikely.

The second possibility is that twice-daily use of an over-the-counter antibacterial mouthwash for 7 d failed to alter the oral microbiome sufficiently to have any “downstream” consequences. This could be because the bacteria found in the oral cavity are extremely resilient, rapidly repopulating themselves after disruption via physical (i.e., toothbrushing) or chemical (i.e., mouthwash) means (Wade 2000). In part, this is because they exist in biofilms on the surfaces of teeth, which are poorly penetrated by cationic agents such as cetylpyridinium (Wade 2000). Nonetheless, cetylpyridinium-containing mouthwashes have been shown to suppress salivary bacterial counts by 40-60% for up to 5 h (Jenkins et al. 1994) and are highly effective at suppressing growth of and/or eradicating bacteria from biofilms *ex vivo* (Roberts et al. 1981; Jenkins et al. 1994; Otten et al. 2011; Dudek-Wicher et al. 2022). Cetylpyridinium-based mouthwash has also been found to markedly blunt increases in salivary and plasma NO_2_^-^ following an oral NO_3_^-^ load (Woessner et al. 2016). Consistent with these prior observations, in the present study there was a significant (P=0.0036) reduction in the percentage of NO_2_^-^ in saliva after 7 d of treatment in the cetylpyridinium group. However, there were no changes in the absolute concentrations of either NO_2_^-^ or NO_3_^-^, suggesting that the effects of cetylpyrdinium on the oral microbiome were somewhat limited.

The third possibility is that cetylpyridinium-containing mouthwash did in fact markedly inhibit NO production via the enterosalivary pathway, but any reduced contribution from this source was simply too small to matter. Consider the following: in the US, normal intake of NO_3_^-^ via the diet is 0.5-1.5 mmol/d (Bryan and Ivy 2015; Piffani-Langley et al. 2024). Absorption of ingested NO_3_^-^ is essentially 100% (Wagner et al. 1984; van Velzen et al. 2008), with the majority being rapidly excreted in the urine and most of the remainder eliminated as other nitrogenous compounds in the feces (Wagner et al. 1984). Prior to its final elimination, though, about 25% of the absorbed NO_3_^-^ is extracted by the salivary glands and secreted into the oral cavity (Spiegelhalder et al. 1976), where roughly 20% is reduced to NO_2_^-^ by oral bacteria (Doel et al. 2005). It has therefore been estimated that under normal dietary conditions only approximately 5% (i.e., 20% of 25%) of ingested NO_3_^-^ is converted to NO_2_^-^ (Bryan and Ivy 2015; Ma et al. 2018). Indeed, we independently arrived at a very similar figure when developing a novel pharmacokinetic model of the combined NO_3_^-^-NO_2_^-^ system (Coggan et al. 2020). Conservatively assuming that all of this newly-formed NO_2_^-^ is ultimately reduced to NO, this implies a maximum rate of NO formation from ingested NO_3_^-^ of 5% of 0.5-1.5 mmol/d, or 0.025-0.075 mmol/d. At the same time, however, in healthy individuals NOS-mediated NO synthesis is approximately 1 mmol/d (Siervo et al. 2011). It is therefore clear that, under normal conditions, the enterosalivary pathway is a relatively minor source of NO, a conclusion supported by previous isotopic tracer research (Rhodes et al. 1995; Castillo et al. 1996). As such, use of antibacterial mouthwash might not be expected to have any marked biological effects unless combined with/compared to ingestion of large amounts of NO_3_^-^ (e.g., Govoni et al. 2008; McDonagh et al. 2015; Woessner et al. 2016) (see below).

Regardless of the above, the present results provide evidence that short-term use of over-the-counter antibacterial mouthwash does not impair muscle strength, speed, or power. As such, these findings are of relevance to both healthy younger individuals interested in athletic performance and older persons or patients with compromised muscle function who might be placed at greater risk of falls if mouthwash use further diminished muscle contractility. The significance of these observations is emphasized by the widespread use of mouthwash in many developed countries (McFarlane et al. 2018; Górska and Górski 2018; Luzzi et al. 2020).

As stated previously, to our knowledge this is the first study to determine the influence of mouthwash use *per se* on exercise capacity. A handful of previous investigations, however, have examined the effects of chlorhexidine-containing mouthwash (which is prescription-only in the US, Canada, and some other countries), sometimes in conjunction with a NO_3_^-^-restricted diet, on other physiological parameters, particularly blood pressure (Kapil et al. 2012; Bondonno et al. 2015; Sundqvist et al. 2016; Ashworth et al. 2019; Bescos et al. 2020). Compared to the present results, these studies have generally found much larger changes in salivary NO_2_^-^ and NO_3_^-^ concentrations, averaging -69% and +80%, respectively, presumably reflecting the greater (more prolonged) antibacterial action of chlorhexidine vs. cetylpyridinium (Wade 2000; Woessner et al. 2016). However, salivary and plasma concentrations of NO_2_^-^ are only weakly correlated (Lumbikanada et al. 2021), and changes in plasma NO_2_^-^ concentration in response to chlorhexidine use have been much more modest (i.e., -19% on average) (Kapil et al. 2012; Bondonno et al. 2015; Sundqvist et al. 2016; Ashworth et al. 2019; Bescos et al. 2020). Furthermore, no changes have been found in plasma cGMP levels (Bondonno et al. 2015; Sundqvist et al. 2016), a sensitive indicator of whole-body NO bioavailability (Kielstein et al. 2004). Perhaps as a result, blood pressure has also been mostly unchanged (Sundqvist et al. 2016; Ashworth et al. 2019; Bescos et al. 2020), with only Kapil et al. (2012) and Bondonno et al. (2015) reporting small, but statistically significant, increases in systolic blood pressure and only Kapil et al. (2015) finding a statistically significant increase in diastolic blood pressure. Again, this could be due to the quantitatively minor contribution of the enterosalivary pathway to NO production under normal circumstances, i.e., at typical levels of dietary NO_3_^-^ intake and in the absence of chronic diseases such as hypertension or heart failure, in which NOS-mediated NO synthesis is reduced (cf. Siervo et al. 2011 for review).

There are a number of limitations to the present study. As implied above, it is possible that we would have obtained different results had we utilized a mouthwash containing chlorhexidine instead of cetylpyridinium, had the participants rinse their mouths more frequently than the recommended two times per day, or had tested the effects of mouthwash in conjunction with a low NO_3_^-^ diet or following NO_3_^-^ ingestion. We might also have found different results had we examined some other exercise outcome, e.g., endurance performance ability, instead of muscle contractile function. Finally, we only measured the concentrations of NO_3_^-^, NO_2_^-^, and cGMP in saliva, such that the impact of the intervention on the concentrations of the metabolites in plasma (or muscle) is unknown. Inclusion of such measurements, however, would not have changed the fact that muscle function was unaltered. Likewise, quantification of the effects of antibacterial mouthwash on oral nitrate reducing capacity would not have altered our other measured outcomes or their interpretation.

In summary, this is the first study to specifically determine the effects of antibacterial mouthwash on exercise capacity, in particular muscle contractile function. Although mouthwash use significantly reduced the relative abundance of NO_2_^-^ in saliva, consistent with inhibition of the enterosalivary pathway, there were no changes in various markers of whole-body NO bioavailability, i.e., the concentrations of NO_3_^-^, NO_2_^-^, and cGMP in saliva and the level of NO in breath, or in maximal muscular strength, speed, or power. Athletes or other individuals for whom muscle contractility may be important (e.g., older persons) seemingly need not be concerned about possible impairments due to use of cetylpyridinium-based mouthwash.

## Data Availability

All data produced in the present study are available upon reasonable request to the authors.

## Abbreviations

ANOVA: Analysis of variance
Ca^2+^: Divalent calcium
cGMP: Cyclic guanyl monophosphate
ELISA: Enzyme-linked immunosorbent assay
NaCl: Sodium chloride
NO_3_^-^: Nitrate
NO_2_^-^: Nitrite
NO: Nitric oxide
PAR-Q: Physical Activity Readiness Questionnaire
Pmax: Maximal knee extensor power
Vmax: Maximal knee extensor velocity

## Author Contributions

EJG, RLH, and ARC conceived and designed the research. EJG, WSZ, AS, and RLH conducted experiments. RLH prepared regulatory documents. EJC, WSZ, AS, and ARC wrote the manuscript. All authors read and approved the manuscript.

## Acknowledgements

EJG was supported by the Diversity Scholars Research Program of the Center for Research and Learning at Indiana University Indianapolis.

